# Estimating the causal effect of genetic liability to prevalent disease on hospital costs using Mendelian Randomization

**DOI:** 10.1101/2020.07.09.20149906

**Authors:** Padraig Dixon, Sean Harrison, William Hollingworth, Neil M Davies, George Davey Smith

## Abstract

**BACKGROUND:** Accurate measurement of the effects of disease status on healthcare cost is important in the pragmatic evaluation of interventions but is complicated by endogeneity biases due to omitted variables and reverse causality. Mendelian Randomization, the use of random perturbations in germline genetic variation as instrumental variables, can avoid these limitations. We report a novel Mendelian Randomization analysis of the causal effect of liability to disease on healthcare costs.

**METHODS:** We used Mendelian Randomization to model the causal impact on inpatient hospital costs of liability to six highly prevalent diseases: asthma, eczema, migraine, coronary heart disease, type 2 diabetes, and major depressive disorder. We identified genetic variants from replicated genome-wide associations studies and estimated their association with inpatient hospital costs using data from UK Biobank, a large prospective cohort study of individuals linked to records of hospital care. We assessed potential violations of the instrumental variable assumptions, particularly the exclusion restriction (i.e. variants affecting costs through alternative paths). We also conducted new genome wide association studies of hospital costs within the UK Biobank cohort as a further “split sample”sensitivity analysis.

**RESULTS:** We analyzed data on 307,032 individuals. Genetic variants explained only a small portion of the variance in each disease phenotype. Liability to coronary heart disease had substantial impacts (mean per person per year increase in costs from allele score Mendelian Randomization models: £712 (95% confidence interval: £238 to £1,186)) on inpatient hospital costs in causal analysis, but other results were imprecise. There was concordance of findings across varieties of sensitivity analyses, including stratification by sex, and those obtained from the split sample analysis.

**CONCLUSION:** A novel Mendelian Randomization analysis of the causal effect of liability to disease on healthcare cost demonstrates that this type of analysis is feasible and informative in this context. There was concordance across data sources and across methods bearing different assumptions. Selection into the relatively healthy UK Biobank cohort and the modest proportion of variance in disease status accounted for by the allele scores reduced the precision of our estimates. We therefore could not exclude the possibility of substantial costs due to these diseases.

**JEL Classification Numbers:** H51, I10, I11, I18,

## 1 Introduction

Liability to disease is influenced by a wide range of factors including environmental, social, economic and biological processes. This complicates attempts to attribute economic outcomes to specific diseases, since measurement error in diagnosis and omitted confounding variables of the association between disease liability and outcome can lead to spurious results. This pervasive endogeneity is problematic for the pragmatic evaluation of medical interventions, which generally require robust estimates of the long term causal effect of disease status on outcomes such as healthcare costs (1-3).

For example, taxes on “sin goods”such as alcohol, cigarettes or sugary beverages are often motivated (amongst other policy rationales) by an anticipated beneficial impact on future morbidity, mortality and healthcare costs (4-6). Even where robust randomized interventional study designs have been implemented, decision-analytic models are usually needed to assess the long-term consequences of policies and interventions (7-14), and will typically assess future costs and other outcomes under different health states (1). If the estimated costs of, for example, coronary heart disease, are biased then robust comparisons of different types of interventions aimed at modifying liability to this disease will not be possible.

The policy relevance of the present work is to develop and validate a method of overcoming the endogeneity that characterizes most or all existing analyses of the long-term impact of disease status on healthcare costs. In turn, our methods may improve the formulation and evaluation of healthcare policies with long-term consequences on healthcare costs. To this end, we report a novel analysis of the causal effect of liability to disease on healthcare costs using Mendelian Randomization (15) which can, in principle, avoid the limitations of existing research designs.

Econometric identification in Mendelian Randomization relies on random perturbations to germline genetic variation that occur at conception. Points of the genome that vary across the population are known as genetic variants. A common form of genetic variation is a single nucleotide polymorphism (SNP) – a change in a single base pair of DNA. At each SNP, everyone has two alleles, one allele that was inherited from the father, one that was inherited from the mother. There is a 50:50 chance of each of the father’s alleles being inherited by the offspring and a 50:50 chance of each of the mother’s alleles being inherited. Therefore, inheritance of genetic variation from parents to offspring is random, conditional on parental genotype.

Some of this variation is known to associate with diseases, behaviours and traits. Those variants known to associate with disease are promising candidates for instrumental variable analysis because conditionally random assignment prior to birth means that these variants are independent of omitted variables that might otherwise confound associations between outcomes and the diseases for which they instrument. Furthermore, estimates using genetic variation are unlikely to be affected by reverse causation since it is generally not possible for the environment to affect germline genetic variation after conception.

We study the costs of liability to six prevalent, chronic diseases: asthma, eczema, migraine, coronary heart disease, type 2 diabetes, and major depressive disorder. If the instrumental variable assumptions (discussed below) hold, then effects liability to disease on these outcomes can be interpreted as causal. Consider, for example, genetic variants that confer susceptibility to coronary heart disease. These variants can be used within the instrumental variables framework to identify the causal effect of coronary heart disease on hospital costs. Thus, genetic variation to higher or lower liability to incident coronary heart disease can be thought of as analogous to allocation to treatment in a randomized controlled trial, in which hospital costs are the outcome.

Below, we describe in more detail each assumption necessary for valid causal inference in the specific context of genetic variants as instrumental variables. We apply these methods to cost data drawn from UK Biobank (16-18), a very large and richly phenotyped prospective cohort study. We also compare the findings of the instrumental variable analysis to conventional multivariable adjusted analyses of the association between disease status and hospital costs. We conclude with a discussion of whether the potential of these methods and data to improve policy formulation and evaluation is likely to be realized.

## 2 Genetic variants as instrumental variables

### 2.1 Instrumental variable assumptions and Mendelian Randomization

Instrumental variables for disease liability are defined by the following assumptions: 1) they associate with liability to the disease of interest, 2) they are independent of all (known and unknown) confounding omitted variables, and 3) they influence the outcome only via disease liability. Instrumental variables have been examined from various perspectives informed by different but largely complementary approaches to causal inference. These particularly include potential outcomes frameworks (19-21) and from the perspective of “Do-calculus”and directed acyclic graphs (22-25).

We unpack the assumptions for valid instrumental variable analysis requirements below for genetic instrumental variables using the language of potential outcomes following von Hinke et al (26) and also illustrate the requirements for valid inference using directed acyclic graphs in supplementary material. Below, we refer to disease status as the treatment variable, and interpret “odds of disease”as a genetic liability (27) toward the specific disease phenotype modelled.

#### RELEVANCE

Valid instruments must exhibit a specific association between instrument and liability to disease. This is sometimes known as the relevance requirement. In terms of potential outcomes, we may express this relationship as the expectation that the value of the treatment variable (disease status in this case) differs when an instrument is “switched on”or has a higher rather than a lower value (in the case of the continuous instruments that we study below), so that the degree of liability to disease differs across values of the instrument. Defining G as the genetic variant (or set of genetic variants), X as the treatment variable (disease status), then G should associate with X: (G<u>∦</u>X).

Instrument relevance is assessed using genome wide association studies (GWASs). These studies examine associations between millions of genetic variants across the genome and phenotypes such as disease status. Humans are diploid, meaning that they have two copies of each chromosome. For a single variant located at a specific part or locus of the genome, an individual may possess no copies of the risk increasing allele, one copy of the risk increasing allele (only on one chromosome) or two copies of the risk increasing allele (on both chromosomes).

For a single variant, there may be two or more alleles (genotypes). The reference allele can be defined as the one that confers the lowest disease risk. The other allele, the risk increasing allele, may confer higher risk of disease. An individual may possess no copies of the risk increasing allele. We can summarize the genotype – the genetic architecture at a particular locus – by simply counting the number of risk increasing alleles (0,1, or 2) that influence disease status. The specific type of variants that we study are single nucleotide polymorphisms (SNPs), which refer to single changes in a nucleotide base pair (adenine (A), thymine (T), cytosine (C), or guanine (G)) at a specific point in the genome across individuals in a population. We use the terms variants and SNPs interchangeably below.

Disease status is considered to be binary. With a binary outcome, and j loci across the genome inspected for possible genetic variation, a GWAS on a binary outcome such as disease status amounts to implementing j separate logistic regressions to assess association. The p-values obtained for each measured association account for multiple testing, reflecting the number of independent genetic variants across the genome. We treated p-values ≤5×10^−8^ as “genome-wide significant”for the purposes of our analysis.

In our main analysis, we use only genetic variants obtained from replicated external GWAS and meet this stringent multiple testing criterion. This reduces the risk of selecting variants that fail the relevance criterion, a problem that affected early examples of Mendelian Randomization in health economics, which sometimes relied on variants drawn from unreplicated “candidate gene”studies (28, 29).

#### INDEPENDENCE

The independence assumption requires the instrument to be independent of potential outcomes and potential disease status, for all possible values of the outcome and disease status. An intuitive interpretation of this assumption is that the instrument is “as good as randomly assigned”. Instrumental variables should therefore be independent of all (known and unknown, measured or unmeasured omitted variables (U): (G<u>∥</u>U)

Genetic instrumental variables are established at conception, and therefore occur before post-natal events and life circumstances. However, while the inheritance of genetic variants from parents to offspring is almost entirely random, the distribution of genetic variants across parents may not be independent of the environment. One means by which the independence assumption may be violated is due to differential ancestry within a population (30). Genetic variants in a population containing several ethnicities with different allele frequencies will associate with any differences between the populations that may be unrelated to the outcome under investigation. For example, allele frequencies and genetic differences in susceptibility to Type 2 diabetes vary by ethnicity (31-35). This mixing of ancestries would lead to spurious associations due to differences between groups that differ in ancestry, rather than the effect only of the genetic variants under investigation.

We minimize this problem by restricting analysis to relatively homogenous European ancestry groups (by means of self-reported ethnicity and conditioning analysis on genetic principal components that reflect variations in ancestry (36) (37)) and by conditioning on study centre. Latent population structure may remain even after these steps an (38, 39) and we consider possible implications in the Discussion section below.

Assortative mating (40) and selection into genetic studies (41) may induce associations between the genetic variants and the treatment variable and outcome that violate the independence assumption. Assortative mating refers to non-random mating; that is, the selection of partners based on particular traits. This may lead to the clustering of particular types of individuals in particular environments; for example, educated people are more likely to marry other educated people. In relation to selection bias, note that conditioning on a consequence (or a collider in the language of directed acyclic graphs) of both the instrument and the outcome can induce an association between the instrument and the outcome even if these are otherwise independent in the population (42, 43). This can lead to bias that can over- or under-state effect sizes (44). Studies necessarily condition on available participants, and if this set of individuals is selected rather than drawn randomly from the population then instrumental variable effect estimates may be biased. Again, we consider the implications of these kinds of potential biases in the Discussion.

#### THE EXCLUSION RESTRICTION

The third assumption is that of exclusion: this requires that the instrument is independent of the outcome (and all potential outcomes) and does not affect the outcome other than via disease status. Genetic variants G should therefore be independent of the outcome Y, conditional on treatment (X – disease status) and all omitted variables (U): (G<u>∥</u>Y | X,U). Establishing? this independence is generally impossible since some omitted variables will not be known or measurable, and disease status may not be measured perfectly.

The two principal means by which genetic variants can violate this assumption are linkage disequilibrium and pleiotropy (45). Linkage disequilibrium refers to a correlation between genetic variants that arises when variants located in close physical proximity to one another tend to be inherited together. Since variants in linkage disequilibrium are not independent, it is possible that correlated variants may influence outcomes other than through the disease of interest in violation of the exclusion restriction.

Pleiotropy describes a situation where a variant affects multiple phenotypes (45-47). There are two types of pleiotropy: vertical and horizontal (48). Vertical pleiotropy refers a SNP that influences one trait, which in turn influences another. Horizontal pleiotropy refers to SNPs that influence traits through independent pathways. Horizontal pleiotropy will violate the exclusion restriction if these other phenotypes affect the outcome of interest other than via the disease of interest.

Consider a variant associated with coronary heart disease that may also influence some other disease (e.g. depression). An analyst using this variant may unwittingly attribute all of the effect of coronary heart disease on cost to that variant, when in truth the reported effect captures the influence of both heart disease and depression on the cost outcome. This amounts to a violation of the exclusion restriction. We consider various methods for modelling and overcoming these violations of the exclusion restriction in the next section.

#### ESTIMATION

The simplest instrumental variable estimator is the ratio of the change in the outcome for a unit increase in the instrument, divided by the change in the treatment variable for a unit increase in the instrument. This instrumental variable estimator recovers the point estimate of the effect and is known as the Wald estimator. It is identical to the familiar two-stage least squares (2SLS) estimator when a single instrument is used. The two-stage estimator predicts disease status (in this case) from a regression of disease status on genetic variants; the second stage involves a regression of the outcome on these predicted values.

In practice we implemented just-identified models where we have one instrument – an allele score or polygenic risk score – using 2SLS, as well as overidentified models which estimate the causal effect using each variant independently. There are two principal reasons for this. The first is that the specific overidentified models that we study allow for a more robust interrogation of the exclusion restriction than is possible using only a 2SLS just-identified model. We describe these advantages below in relation to the specific estimators that we implement. The second is that ratio-type estimators particularly lend themselves to the analysis of summary data that characterize the association of genetic variants with disease and outcome at the level of the variant rather than at the level of the individual person.

These summary data can be readily analyzed using two-sample instrumental variable methods (49), in which instrument-outcome and instrument-disease status are ascertained in distinct samples. This permits analysis of summary data drawn from large numbers of individuals provided that the populations of individuals in each study are similar (50, 51).

We point identify our estimators by assuming that the instrument has a monotonic effect on the treatment variable (52, 53). This assumes that, in a hypothetical experiment, replacing an allele with no effect on disease liability with a liability-increasing allele would either increase liability or leave it unchanged. This implies that, across our study population, the direction of effect from modulating the value of our instrumental variables will be in the same direction for everyone.

Monotonicity may have biological plausibility for genetic instruments (54), but this is generally difficult or impossible to test. Under our maintained assumption that monotonicity applies, our estimates represent local average treatment effects – these effects are local to those individuals whose liability to disease is altered by the instrument. This group of individuals may comprise everyone in our sample if liability to disease is continuous.

We also examine whether our instruments are weak. A weak instrument explains only a small portion of the variance in disease status. Intuitively, when the denominator of the ratio estimator is close to zero, the ratio becomes unstable and its variance becomes large. We examined first stage F-statistics from 2SLS allele score models to assess whether weak instruments were likely to be problematic.

### 2.2 Sensitivity analysis

Our main sensitivity analyses comprised various tests designed to assess possible consequences arising from violations of the exclusion restriction, but we also considered biases arising from differences between the samples used in the two-sample analysis.

Sensitivity analysis directed at potential violations of the exclusion restriction relies on techniques influenced by meta-analysis (55), the motivation for which stems both from the independence of variants as well as from interpreting conditional random allocation of genetic variation at conception as analogous to a within-family randomized controlled trial in which the individual is allocated to higher or lower susceptibility to disease. When many such variants are available, we may proceed, as in a conventional meta-analysis, to summarize their effects on the outcome in a manner that is at least as informative as the scrutiny of any one variant. A variety of these techniques are applied, each with different assumptions. Similarity or concordance between the results in spite of different assumptions provides a degree of reassurance that the same causal effect is being identified, and that gross biases specific to one form of analysis are not influencing the results.

The starting point for this analysis of individual variants is to implement an inverse variance weighted (IVW) random effects meta-analysis. This weights the effect sizes (i.e. the regression betas) by the inverse of the variance of the estimated associations between the variants and the outcome. Since summary causal effects are determined only by the ratio of the effect of variants on the outcome to effects of variants on disease liability, the exclusion restriction requires that there should be no effect on the outcome for any SNP that does not also affect the treatment variable.

A random-effects meta-analysis allows some or all variants to have pleiotropic effects in violation of the exclusion restriction, but assumes that the effect of this pleiotropy “balances out”so that pleiotropy that increases the causal effect estimate is matched by pleiotropy that reduces the causal effect estimate. Thus, the effect size is estimated without bias, although with a greater standard error than would be the case without variation induced by pleiotropy.

A natural test of the assumptions of this model (embodying the assumption of no directional pleiotropy) is to compare this IVW effect estimate with the effect estimate for each individual variant. If this difference is large (in a sense defined below), then heterogeneity of effect may be present. Heterogeneity may indicate horizontal pleiotropy, since one mechanism to generate heterogeneity of this type would be the influence of a variant on the outcome through multiple independent channels.

This can be tested formally using Cochran’s Q statistic, which measures heterogeneity by comparing the squared difference to the critical values of a chi-squared distribution:

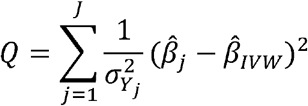

Here, we assume up to J variants, and measure effect estimates for the jth variant as 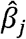 and the overall inverse variance weighted estimate as 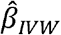, and measure the inverse variance weighting of the estimate using 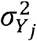 which is the variance from the SNP-outcome association. The Q statistic is identical to the Sargan (56) test for overidentification when all relationships (SNP-treatment and SNP-outcome) are measured in the same sample (57).

We estimate the Q statistic for all diseases and implemented pleiotropy-robust methods if heterogeneity was indicated. These were of two broad types – the first type of sensitivity analysis assumes that some or all variants violate the exclusion restriction but are nevertheless retained in the analysis. The second type of approach comprises a principled approach to variant outlier detection and removal, and analysis may be applied to a smaller, restricted set of variants. We consider each approach in turn; slightly more formal derivations are available in Dixon et al (58) while supplementary material provides an intuitive explanation using simple graphical examples.

We start with estimators of the first type. Median-based estimators (59) assume that the median estimate, constructed by forming an empirical density of all estimates, is unbiased (i.e. has no horizontally pleiotropic effects). Excluding pleiotropic variants will mean that they offer no weight to the calculation of the overall summary effect. Thus, if at least half of the weighted variants do not have horizontally pleiotropic effects, this method will be asymptotically unbiased. We implement a penalized weighted median estimator, in which variant-specific effects are weighted by the precision with which they are estimated and penalized (or “down-weighted”) for contributing heterogeneity to the Q statistic.

A related method makes an assumption that modal values of individual variants are not in violation of the exclusion restriction (60). This permits some SNPs, possibly most SNPs, to be pleiotropic and therefore in violation of the exclusion restriction, provided that the modal variant is, or modal set of variants are, unbiased. Again, we implement a weighted version of this estimator to account for the precision with which variants are estimated, and we also define an arbitrary bandwidth parameter to identify the modal group.

We also implement MR Egger regression (61). The IVW, penalized weighted median and weighted mode constrain the intercept to be zero so that the regression line passes through the origin. MR Egger does not constrain this intercept to be zero. Instead, it is included and can be interpreted as the average pleiotropic effect from all variants. An intercept that is distinguishable from zero indicates the presence of pleiotropy, and the causal effect estimate from MR Egger regression is unbiased, conditional on pleiotropy (if present). This method relies on an assumption about independence between the direct pleiotropic effect of variants on the outcome (other than through liability to disease) and instrument strength – for further details see Bowden et al (61) and Burgess and Thompson (62).

We also implement an approach, known as Radial Mendelian Randomization (63), to automate the identification of outlying variants. In a scatter plot of variants (see illustrations in the supplementary material) that plots the SNP-outcome association (y-axis) against the SNP-disease status association (x-axis), the absolute vertical distance between a variant represented by a scatter point and the fitted IVW line is equivalent to the square root of that variant’s contribution to Cochran’s Q heterogeneity statistic. This distance can therefore be used to identify outlying variants that contribute relatively high amounts of heterogeneity by using a leave-one-out analysis of all variants. We implement all estimators described above on exclusion of variants identified by this process and compare results to models using the full set of variants.

Finally, to complement the two-sample approach described above, we also performed a one-sample analysis using data only from UK Biobank. One concern with two-sample estimators is that the populations in each sample should not overlap (that is, not have the same individuals) to avoid inducing a correlation between SNP-disease and SNP-outcome associations and biasing estimates towards the non-IV estimate. However, the two samples should be drawn from the same population, even if the set of individuals in each sample is disjoint. The simple rationale for this is to avoid bias caused by associations estimated on samples that differ in the types of individuals included. Similarity may be assessed in relation to distributions of age and sex, similarity of association between SNPs and the phenotype, and the mean and variance of the phenotype distribution (50).

We split UK Biobank into two random, non-overlapping samples for this sensitivity analysis. This approach eliminates any biases caused by sample heterogeneity between the UK Biobank cohort and external GWAS source data. We conducted a de novo GWAS for each of the six diseases on each split sample. The result of each GWAS was used to identify SNPs for the alternative sample. We estimated just-identified allele score models using each sample, and then used fixed effect meta-analysis on these separate results to give a single estimate.

#### 2.3 Issues of interpretation

The definition of disease reflects a binary classification of individuals as cases (those identified as having the condition) and controls (those without). The interpretation of our results differs according to the type of instrumental variable models used – those that use a single allele score and those that use each SNP as individual instruments.

The interpretation of the allele score instrumental variable models reflect the genetic liability of changing from control to case across the population. Put differently, these models may be interpreted as the average per person change in hospital costs caused by a (genetically influenced) change in case status in this population. These models are estimated using 2SLS. Denoting individuals *i*=*i…N, k*=*1…K* instruments (G=*g_1_*,…,*g_k_*) and disease X=*x*_*1*_,*…,x*_*6*_ the first stage of a 2SLS model is:

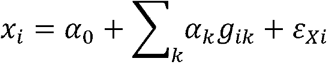

This yields predicted values of disease status: 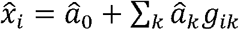, which enter the second stage regression for outcome Y=y_1_,…,y_i_:

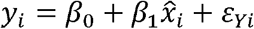

The *β*_1_ coefficient represents the estimated effect of a change in disease status.

The interpretation of the models that use summary data is different. An intuitive way of understanding this difference is to consider a simple ratio estimator for instrumental variables.

The instrumental variable point estimate may be obtained as the change in the outcome (Y) for a unit increase in the instrument (G), scaled by the change in disease status (as the treatment variable) for a unit increase in G:

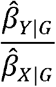

The interpretation of Mendelian Randomization models using each SNP as an individual instrument therefore reflects the measurement scale used in the source GWAS, in contrast to the just-identified allele score models estimated using 2SLS. Since the GWASs we used analyzed outcomes using logistic regression, our over-identified models using each SNP as an instrument therefore measure the ratio of the effect of the change in costs per unit change in the log-odds of disease status.

This type of association can be difficult to interpret in terms of causal effects since a unit change in a binary disease status variable will be on the log-odds scale. A unit change is therefore exp(1), corresponding to a 2.72-fold (since the scale is multiplicative) change in the odds of the treatment variable. Following Burgess and Labrecque (64), we re-express associations estimated from models relying on individual variant data to aid interpretation. We interpret “odds of the treatment variable”as a genetic liability (27) toward the specific disease phenotype modelled. We calculate causal effects in terms of a doubling in genetic liability (ln2) and a 10% increase (ln 1.1). In the main text we report estimates according to a doubling in genetic liability; inverse variance weighted estimates of the effect of a 10% relative increase are reported in the supplementary material.

Estimates on the relative scale may therefore be interpreted as, for example, the causal effect on inpatient hospital costs of a doubling in genetic liability to coronary heart disease. Note that this is a relative rather than absolute increase, where the relative increase can be understood as an increase from possessing some risk-increasing alleles to more risk-increasing alleles.

### 2.4 Data

The principal source of our data is the UK Biobank, a large population-based prospective cohort that recruited over 500,000 adults aged between 39 and 71 over the period 2006-2010 (13–15). Participants provided data on demographic features such as date of birth, sex as well as a wide variety of phenotypic data. Approximately 9 million individuals registered with the National Health Service were invited to participate (16, 17). The participating cohort of over 500,000 individuals is generally “healthier and wealthier”(16, 41, 65) than the general UK population. We discuss some of the consequences of this feature of the UK Biobank cohort below.

These data were linked to records of inpatient hospital care. We ascertained disease status (other than depression – see below) from self-report at recruitment interview (including of the general form “Has a doctor ever told you that you have had any of the conditions below?”) and from a review of ICD-9 and ICD-10 codes coded against inpatient hospital encounters that took place before the baseline visit. The diabetes cases were only those known to have Type 2 rather than Type 1 diabetes (66).

The depression cases were defined (as in Tyrrell et al (67) and in Harrison et al (66)) as participants who self-reported seeing a doctor for nerves, anxiety or depression for a duration of at least two weeks, or who had ICD-9 or ICD-10 codes for depression. Only ten recruitment centres asked questions related to depression, and this reduced the size of the available sample (see Results below) relative to the other conditions.

We studied non-cancer diseases with known genetic determinants, for which prevalence was at least 2% in the UK Biobank, and which accounted for at least 100 disability-adjusted life years lost per 100,000 adults in the UK (68). Diseases with less than three genome-wide significant SNPs were excluded from consideration, which resulted in the exclusion of osteoarthritis. The final set of diseases resulted in analysis sample comprising the following six diseases: asthma, eczema, migraine, coronary heart disease, type 2 diabetes and depression.

Our outcome measure is inpatient hospital costs, for which a median of 6.1 years of follow-up was available. Episodes of hospital care were converted into healthcare resource groups, which are summary codings that group together similar types of care received. These codings account for diagnosis, procedures and complexities such as complications and comorbidities. Healthcare resource groups were calculated using an NHS algorithm (69) used for the remuneration of care provided in the public healthcare system. The healthcare resource groups so created were linked to published data on the per unit cost of each such group, allowing a per patient, per year hospital cost figure to be generated. Further details on this process are available in Dixon et al, (70, 71).

We examined previous GWAS for data on variants associated with disease at genome-wide significance (P ≤ 5 × 10^−8^). The source GWAS data for each condition was as follows: asthma (Moffatt et al, (72)), coronary heart disease (Schunkert et al, (73)), major depressive disorder (Wray et al, (74)), Type 2 diabetes (Morris et al, (75)), eczema (Pasternoster et al, (76)), migraine (Gormley et al, (77)). The Wray et al (74) GWAS included individuals from the pilot sample of UK Biobank, and we excluded these individuals in the two-sample analysis to avoid overlap between the GWAS and analysis samples.

We cross-checked genome-wide significant variants against those measured in the UK Biobank cohort. In some cases we did not locate in UK Biobank the precise variant used in the external GWAS – the difference between the precise variant used typically arises because of the different technologies used to genotype different cohorts, and the areas of the genome searched. Where the genetic variant was not available in UK Biobank, we used a substitute variant with a high (R^2^>0.6) degree of correlation with the missing variant, or if no substitute could be found, we excluded the missing variant.

We also created allele scores for each condition, also sometimes known as polygenic risk scores. The number of disease-increasing alleles was counted for each individual, then multiplied by the effect size, determined in the respective GWAS, for each allele a weighted per-allele effect was summed. This gives a weighted allele score, where the weights reflect the effect size of each allele.

To avoid bias induced by differential ancestry,. For all models, we condition on sex, age and the first forty genetic principal components. The analysis sample was restricted to individuals of white British or closely related ancestry (determined by self-report and by examination of genetic principal components) to avoid bias from differential ancestry. Analyses were conducted in R (including with MR Base package (78)) and in Stata version 15.1 (StataCorp, College Station, Texas). Analysis code is available at github.com/pdixon-econ

## 3 Results

### 3.1 Introduction

Up to 307,032 individuals were available for inclusion in the inferential analysis, of whom 54% were female (n=164,985). Mean age at recruitment in this sample was 57 years (SD = 8.0 years). The actual numbers available for the analysis of each specific condition were slightly lower than the overall total given the completeness of data on self-report of disease history at baseline and information available to identify cases in linked hospital records.

Multivariable models, estimated using linear regression and adjusted for age, sex and study centre but without any genetic information, indicated substantial impacts on inpatient hospital costs of most diseases. This is summarized in Table 1. The coefficients represent the association of disease status with hospital costs.

**Table 1.**
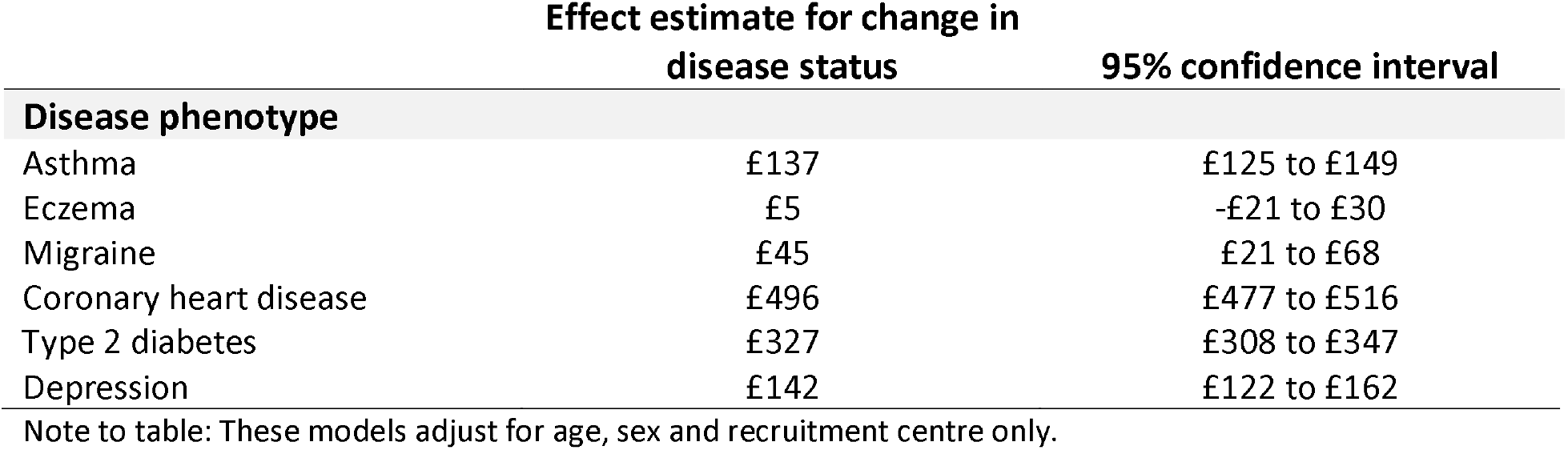
Multivariable estimates of association between disease status and annual inpatient hospital costs.

With the exception of eczema, all disease phenotypes are associated with material impacts on annual per-patient inpatient hospital costs. We emphasize that these models are minimally adjusted (accounting only for age, sex and centre) but the scale of some of these effects (particularly for coronary heart disease) is large relative to median (£88) and mean (£479) annual per-patient inpatient hospital costs.

These estimates are likely to be confounded in various ways, particularly by omitted variables that influence both hospital costs and disease status. This possibility of confounding constitutes our motivation for considering more robust causal methods using genetic liability to disease, to which we now turn.

We begin by considering the proportion of variance in the disease phenotype explained by respective allele scores (measured by pseudo-R-squared statistics obtained from unconditional logistic regressions) summarized in Table 2.

**Table 2.** Cases, number of SNPs and strength of instruments.

| **Disease phenotype** | **N** | **N of cases (%)** | **N of SNPs** | **% of variance explained by allele score in disease status** | **F-statistic from first stage of 2SLS allele score model** |
| --- | --- | --- | --- | --- | --- |
| Asthma | 306,245 | 39,159 (12.8%) | 8 | 0.53% | 1,242 |
| Eczema | 306,245 | 8,017 (2.6%) | 12 | 0.37% | 270 |
| Migraine | 306,245 | 9,782 (3.2%) | 33 | 0.54% | 484 |
| Coronary heart disease | 306,245 | 14,261 (4.7%) | 15 | 0.43% | 520 |
| Type 2 diabetes | 304,885 | 13,444 (4.4%) | 13 | 1.1% | 1,274 |
| Depression | 92,830 | 18,760 (20.0%) | 36 | 0.10% | 93 |

Notes to table: Depression is reported for fewer individuals than other phenotypes because not all Biobank recruitment centres asked individuals about depression and also because the principal source for these data (Wray et al (74)) included the UK Biobank pilot sample, information from which we have excluded to ensure independence of GWAS and analysis samples. Diabetes cases were limited only to those with Type 2 rather than Type 1 cases.

Table 2 makes clear that the proportion of variance explained by available SNPs is modest, being less than 1% in all cases. However, the associated instrumental variables are unlikely to be affected by weak instrument bias given that the p-values associated with the first stage F statistic were <0.0001 for all diseases. Allele scores for each condition were not strongly related to sex for any condition, and only the type 2 diabetes score showed possible evidence of an association with age at recruitment (p-value=0.04).

### 3.2 Main results

Table 3 summarises the causal effect estimates and associated 95% confidence intervals for all six disease phenotypes as estimated using just-identified allele score 2SLS models. The betas in Table 3 reflect inpatient hospital costs per person per year of a (genetically influenced) change in case status from control to case.

**Table 3.** 2SLS allele score estimates.

| Disease phenotype | Beta | 95% confidence interval |
| --- | --- | --- |
| Asthma | -£30 | -£222 to £161 |
| Eczema | £621 | -£239 to £1,482 |
| Migraine | £786 | £199 to £1,373 |
| Coronary heart disease | £712 | £238 to £1,186 |
| Type 2 diabetes | £173 | -£132 to £479 |
| Depression | -£348 | -£997 to £300 |

**Table 4.** Cochran’s Q statistic and heterogeneity by phenotype.

|  | N of SNPs | Q statistic | Q p-value |
| --- | --- | --- | --- |
| Disease phenotype |  |  |  |
| Asthma | 8 | 8.6 | 0.28 |
| Eczema | 12 | 16.9 | 0.11 |
| Migraine | 33 | 38.2 | 0.09 |
| Coronary heart disease | 15 | 26.7 | 0.02 |
| Type 2 diabetes | 13 | 19.3 | 0.02 |
| Major depressive disorder | 36 | 48.7 | 0.02 |

Estimates for asthma, eczema, Type 2 diabetes, and depression were consistent with the null. Migraine and coronary heart disease were consistent with a large positive effect on costs. It is notable that confidence intervals are wide in all cases, reflecting the modest proportion of variance in disease status that is explained by each respective score.

Stratification of the 2SLS allele score models by sex did not reveal any material differences to unstratified 2SLS models (Figure 1), although it is notable that effects for migraine may be most influenced by females cases, and effects for coronary heart disease by male cases.

**Figure 1.**
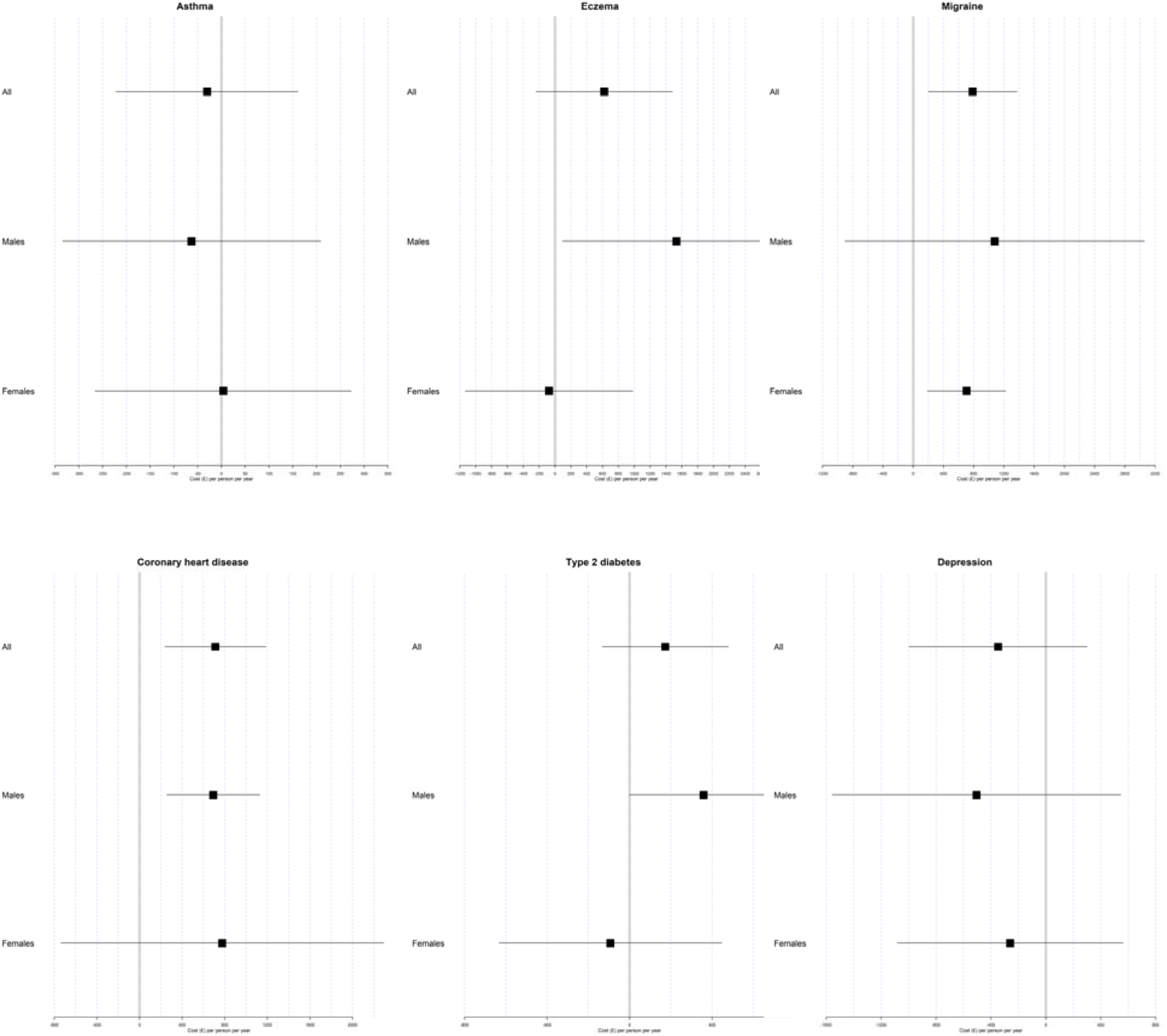
2SLS allele score estimates stratified by sex.

Cochran’s Q statistic revealed some evidence of heterogeneity for all diseases, although the evidence for asthma is relatively weak.

Inspection of forest plots measuring the causal effect of each SNP for each condition reveals this heterogeneity. In Figure 2, heterogeneity is indicated by asymmetry about the lines of null effect. Some differences in the magnitude of effect are expected due to sampling variation, but more substantial heterogeneity is apparent.

**Figure 2.**
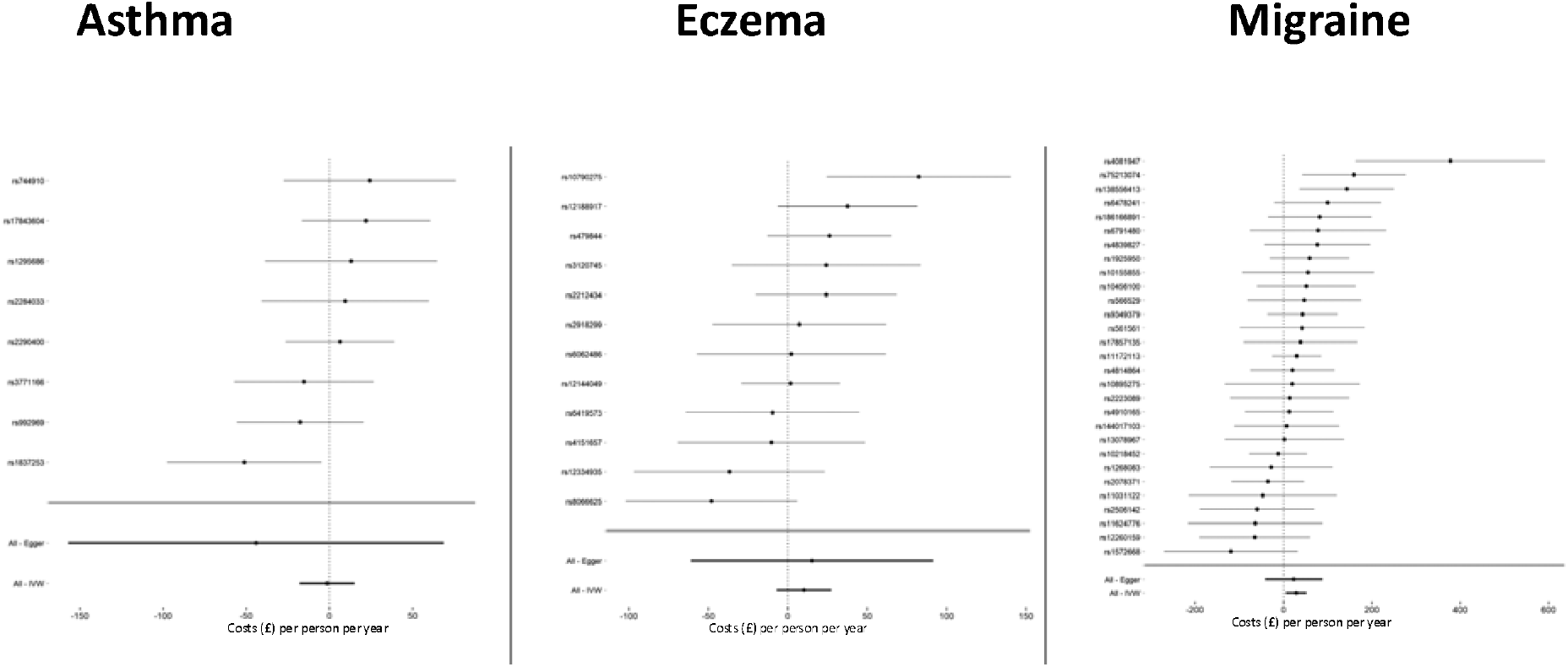
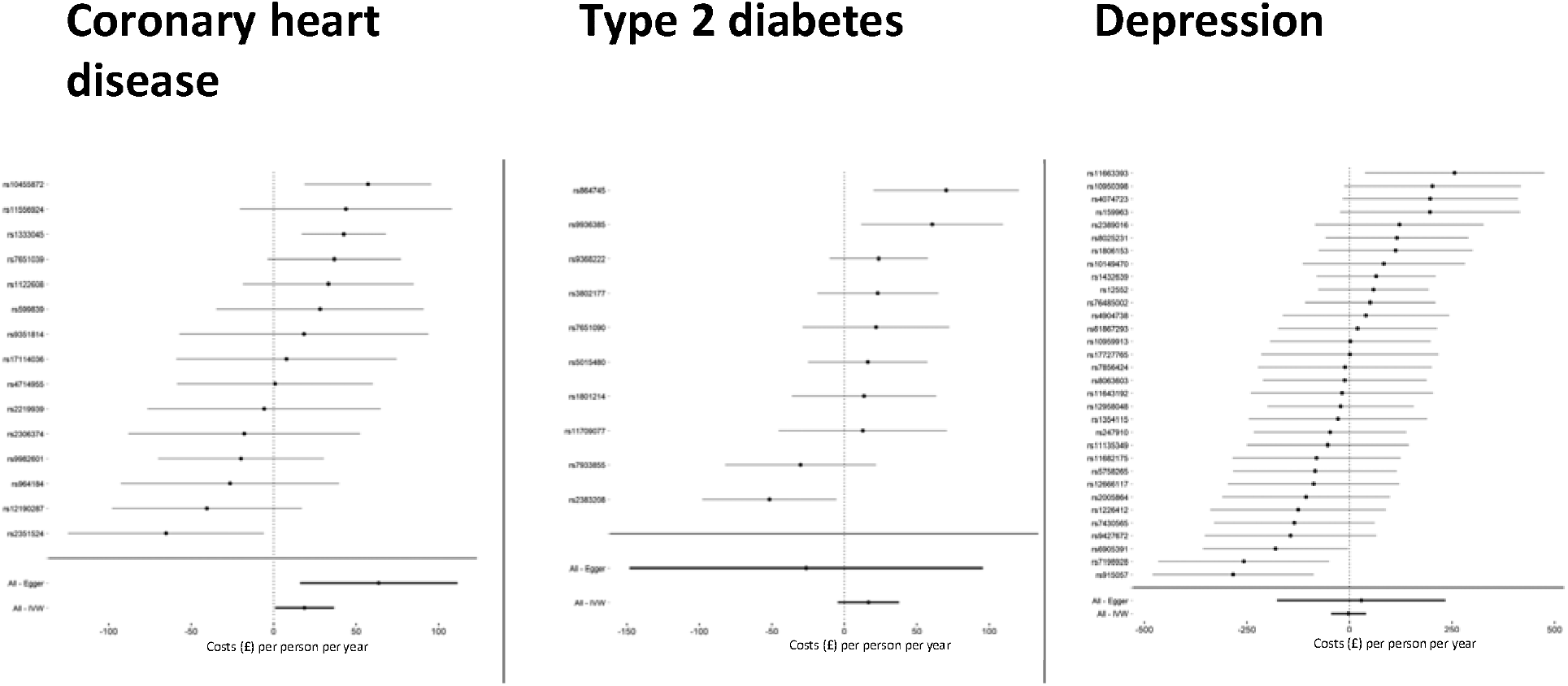
Forest plot of genome-wide significant SNPs for all conditions (scaled to reflect a 100% relative increase in genetic liability)

To avoid imposing a sharp cut-off for “significant”evidence of heterogeneity, we conservatively apply various forms of pleiotropy-robust sensitivity analysis to all phenotypes. Consistency among estimators, and between the types of sensitivity analysis, offers some reassurance that similar causal effects are identified, irrespective of the type of assumptions made regarding violations of the exclusion restriction. Supplementary material presents scatter plots for all variant and conditions.

Figure 3 summarises the results of for all conditions using all pleiotropy-robust methods. These forest plots contain point estimates and 95% confidence intervals on a condition-by-condition basis for the six diseases. These include both the full set of SNPs and restricted SNPs following removal of outliers identified by the radial methods described above.

**Figure 3.**
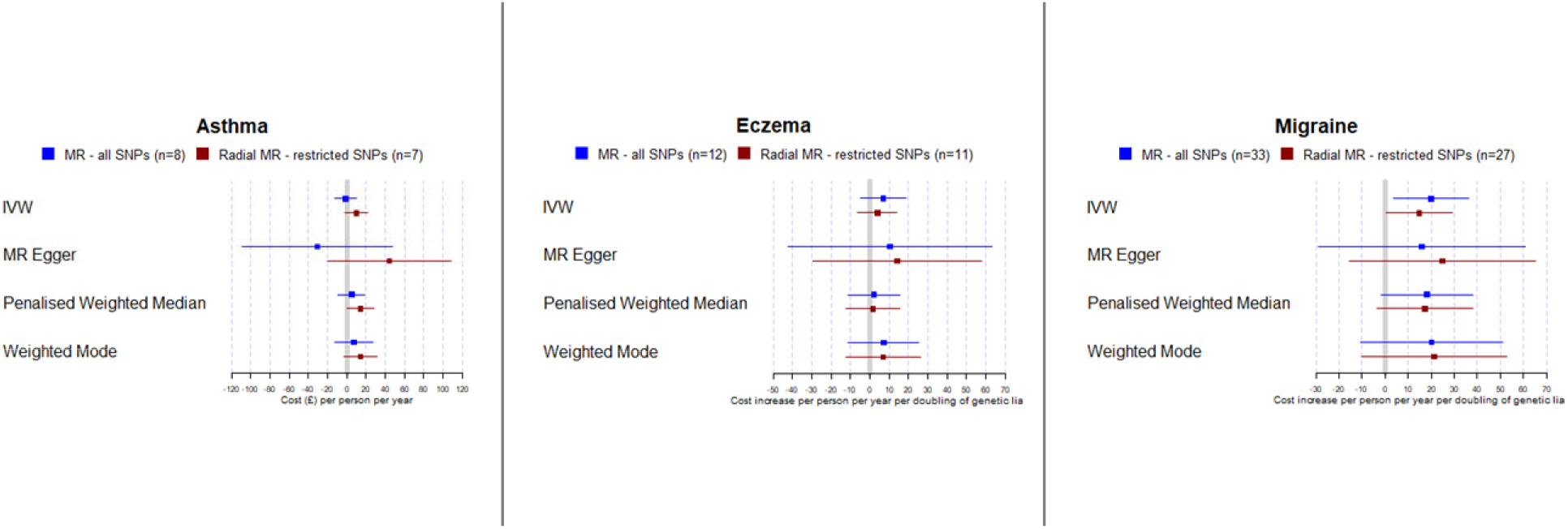
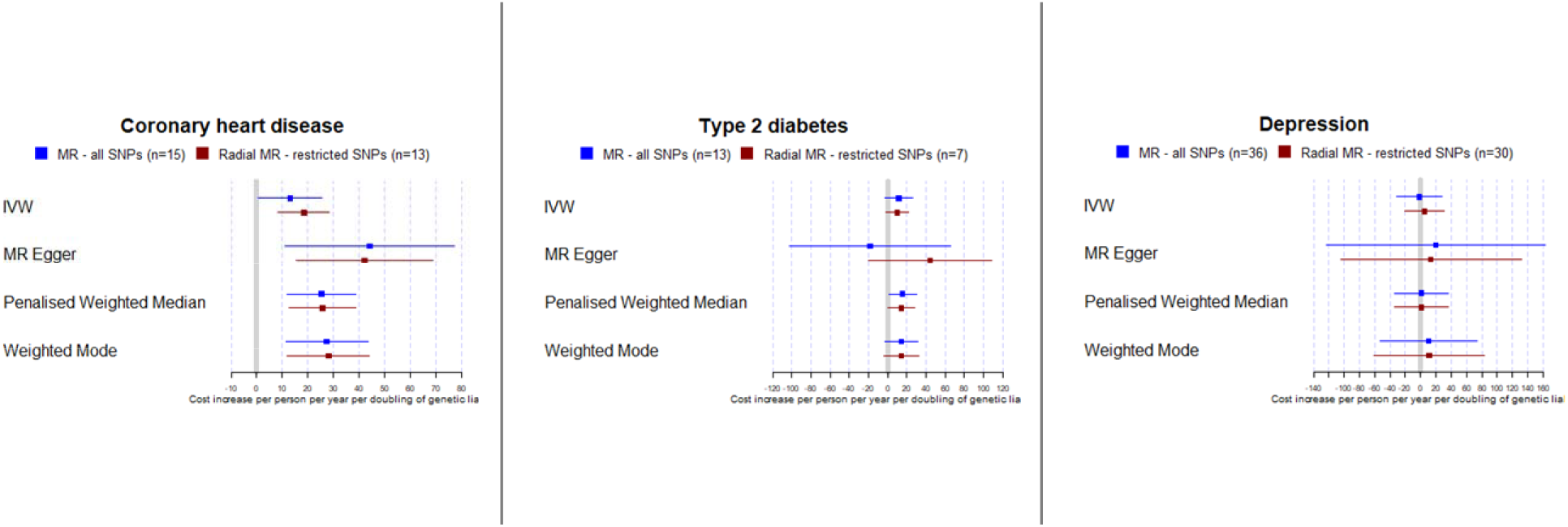
Comparison of summary estimators. Note to figure: IVW – inverse variance weighted.

It is notable that effect estimates are similar within each disease regardless of whether some or all SNPs are included in conventional or radial Mendelian Randomization models. These estimates reflect per person, per year inpatient hospital costs on a relative scale: all estimators for all conditions imply modest effect sizes per doubling of relative genetic liability. The estimates of coronary heart disease are again more precise than for other disease.

The split sample results for the allele score models are presented in Table 5. We did not identify sufficient genome-wide significant SNPs to calculate allele scores for depression.

**Table 5.** 2SLS allele score estimates from split sample analysis.

| Disease phenotype | Beta | 95% confidence interval |
| --- | --- | --- |
| Asthma | £87 | -£26 to £199 |
| Eczema | £510 | -£50 to £1,069 |
| Migraine | £116 | -£610 to £841 |
| Coronary heart disease | £1,123 | £672 to £1,573 |
| Type 2 diabetes | £222 | -£25 to £470 |

Comparisons between the results of the split sample with those of the main analysis are subject to a number of qualifications, particularly because these models comprise different sets of individuals. Confidence intervals for each phenotype overlaps in each analysis. Asthma has the smallest effect size in both the main analysis and the split sample analysis, while coronary heart disease has the largest effect size in the split sample analysis but only the second largest in the main analysis. We cannot calculate a Cochran Q value for the meta-analyzed split sample data given the differences in the SNPs identified as genome-wide significant in each sample. Overall, there is little evidence of any gross difference between the split sample allele score results and those of the primary analysis.

## 4 Discussion

Our analysis was motivated by the important observation that the long-term consequences of various types of intervention can only be evaluated if the cost data used in those evaluations is unbiased. Confounding by omitted variables that jointly influence disease status and healthcare costs is likely to be the principal source of bias that affects existing methods for understanding these relationships. If the instrumental variable assumptions hold, our methods will produce unbiased estimates of the cost of increasing genetic liability to disease.

We used the largest available genome-wide association studies to identify SNPs related to six prevalent chronic diseases that are each associated with substantial morbidity in the general population. Effect sizes estimated using Mendelian Randomization models of genetic liability to disease were larger than those obtained from simple linear models and were roughly concordant across different types of sensitivity analysis but were substantially more imprecise. The similarity of results obtained from different Mendelian randomization estimators offers some reassurance that the same causal effect was being identified even when assumptions regarding violations of the instrumental variable assumptions differed.

We emphasize that our causal estimates are most appropriately interpreted as relating to liability to disease. It is possible in some cases that effect of liability on healthcare costs will be most relevant to a particular use of these findings; in other cases it may be less so. Nevertheless, there is an important direct link between disease liability and ultimate disease status. Interventions that reduce liability to the six diseases we study may reduce healthcare costs; for example, smoking cessation interventions may ultimately reduce the costs associated with liability to incident coronary heart disease, even in those individuals who do not develop that disease.

Estimates were imprecise when considering either the just-identified allele score models, or the summary sensitivity analyses that rely on over-identified models using data on many individual genetic variants (including over-identified models that removed outliers), and in both the main analysis and the split-sample analysis. There was strong evidence of a substantial and economically meaningful impact of coronary heart disease status on costs, but much less clear evidence for the other disease phenotypes. No differences were apparent when 2SLS allele models were stratified by sex.

Confidence intervals were wide for most diseases and were consistent with both substantial positive and negative effects. This may be because of the modest proportion of variation in disease status explained by available SNPs (79). This contrasts with other work using continuous exposures in relation to hospital costs, for which statistical power will be greater, such as body mass index (58). Further GWASs to increase the number of SNPs and proportion of variance explained in these disease phenotypes will increase the power of this analysis, as will even larger data resources that link to resource use and cost data. Future studies with improved precision will clarify whether Mendelian randomization estimates do indeed imply larger effect sizes than those from the conventional multivariable adjusted models.

A second mechanism that may have affected the size and precision of effect estimates is selection (41) into UK Biobank. This cohort is more educated, wealthier and has a more favourable risk profile for adverse health outcomes than the general UK population from which the study population was drawn. We cannot quantify the extent of this bias without knowledge of selection mechanism itself. Analyses of selection bias in (41, 42) in Mendelian Randomization studies suggest that other sources of bias (such as pleiotropy) will be more consequential, although relative magnitudes of bias will depend to some extent on study context.

A further source of potential bias is geographic stratification in allele distributions that is not removed by conditioning on genetic principal components (38, 80). This can lead to confounding associated with environmental factors that differ by region. Again, the precise extent of this influence on our results is unknown, but merits consideration in their interpretation. Recent evidence on these magnitudes suggest that bias is likely to be modest given recent evidence indicating an absence of geographic structure in allele scores for all diseases other than coronary heart disease (81).

The random allocation of genotypes that is the foundation of Mendelian Randomization is conditional on parental genotype, meaning that the randomization occurs within the family unit (15, 82-86). Dynastic effects (29) arise when the expression of the genetic variants in the parents affects disease status in the offspring independently of the child’s genome. For example, it could be the case that genetic associations of asthma are influenced by parental smoking behaviour. Causal analysis relying on these SNPs would wrongly attribute causal effects to the asthma SNPs alone, when in truth the effects of the environment were also material to the outcome.

Within-family studies would offer the best source of evidence on possible dynastic effects. However, there are far fewer available samples of siblings meaning that within-family estimates are unlikely to be informative because statistical power would be much reduced relative to the analysis of unrelated individuals that we report for our main analysis (87) (82). Likewise, data on family trios (parents and offspring) would help assess whether assortative mating is likely to bias (40) our Mendelian Randomization analyses of unrelated individuals. In practice, biases of this type may be less plausible for the types of diseases we study compared to traits such as BMI (88, 89) and education (90), for which some evidence of these types of effect has been identified.

Our instrumental variable models estimated local average treatment effects of random allocation to lifelong liability to each of six diseases. Note also that Mendelian Randomization analyses may not comply with the stable unit treatment value assumption (SUTVA) of causal inference (19), which requires that there is a single version of the treatment analyzed. Treatment, in the present context, refers to liability to disease, and the assumption is violated because we do not know if a (hypothetical) manipulation of in individual’s genome to alter liability has an identical effect to other means of managing disease liability. For instance, various behavioral and socioeconomic factors are implicated alongside genetic influences in the etiology of coronary heart disease (91, 92). Our results estimating the causal effect form SNPs implicated in this disease do not necessarily correspond to the results that might be obtained from a hypothetical manipulation of these other risk factors through, for example, taking statins to manage the risk of heart disease.

## 5 Conclusion

We report a novel Mendelian Randomization analysis of the causal effect of liability to disease on healthcare cost. Our analysis demonstrates that this type of analysis is both feasible and informative in this context. Results were consistent with both large and small effects of disease on inpatient hospital costs. There was concordance across data sources and across methods bearing different assumptions. The modest precision of available data indicating genetic liability to prevalent long-term health conditions, and selection into the relatively healthy UK Biobank cohort, is likely to explain the imprecision, suggesting the absence of evidence rather than evidence of absence. These considerations will challenge future work research using data from these and similar cohorts, but nevertheless indicate the rich potential for future work.

## Data Availability

The principal source of data used in this study is the UK Biobank, the terms of access of which do not permit data sharing by third parties. However, data used in this study are available via application to UK Biobank. Other data used int he study are drawn from publicly available genome-wide association summary statistics, the sources for which are cited in the main text.

## Acknowledgments

The authors are grateful to comments on this paper from Peter C. Smith and other participants at the Health Economics Study Group in Newcastle, January 2020. This research has been conducted using the UK Biobank Resource under Application Number 29294.

